# Childhood maltreatment and subsequent risk of hospitalization or death due to COVID-19: a cohort study in the UK Biobank

**DOI:** 10.1101/2023.09.12.23295354

**Authors:** Yue Wang, Fenfen Ge, Thor Aspelund, Helga Ask, Arna Hauksdóttir, Kejia Hu, Jóhanna Jakobsdóttir, Helga Zoega, Qing Shen, Heather C Whalley, Ole Birger Vesterager Pedersen, Kelli Lehto, Ole A Andreassen, Fang Fang, Huan Song, Unnur A Valdimarsdóttir

**Affiliations:** Center of Public Health Sciences, Faculty of Medicine, University of Iceland, Reykjavík, Iceland; Institute of Environmental Medicine, Karolinska Institutet, Stockholm, Sweden; Department of Mental Disorders, Norwegian Institute of Public Health, Oslo, Norway; Department of Psychology, University of Oslo, Oslo, Norway; School of Population Health, Faculty of Medicine and Health, UNSW Sydney, Sydney, Australia; Clinical Research Center for Mental Disorders, Shanghai Pudong New Area Mental Health Center, Tongji University School of Medicine, Shanghai, China; Institute for Advanced Study, Tongji University, Shanghai, China; Centre for Clinical Brain Sciences, University of Edinburgh, UK; Generation Scotland, Institute of Genetics and Cancer, University of Edinburgh, UK; Department of Clinical Medicine, Faculty of Health and Medical Sciences, University of Copenhagen, Copenhagen, Denmark; Department of Clinical Immunology, Zealand University Hospital, Denmark; Estonian Genome Centre, Institute of Genomics, University of Tartu, Tartu, Estonia; NORMENT Centre, Institute of Clinical Medicine, University of Oslo, Oslo, Norway; NORMENT Centre, Division of Mental Health and Addiction, Oslo University Hospital, Oslo, Norway; West China Biomedical Big Data Center, West China Hospital, Sichuan University, Chengdu, Sichuan, China; Med-X Center for Informatics, Sichuan University, Chengdu, Sichuan, China; Department of Epidemiology, Harvard T H Chan School of Public Health, Boston, MA, USA

## Abstract

Childhood maltreatment has been associated with some infection-related outcomes, yet its potential role in severe COVID-19 outcomes has not been addressed. Therefore, leveraging longitudinal data from the population-based UK Biobank (N=151,427), our study aimed to explore the association between childhood maltreatment and severe COVID-19 outcomes (i.e., hospitalization or death due to COVID-19) and its underlying mechanisms. Our results *suggest* that childhood maltreatment, particularly physical neglect, is associated with a 54.0% increased risk of severe COVID-19 outcomes (i.e., hospitalization or death due to COVID-19), which was not modified by genetic predisposition to severe COVID-19 outcomes. *We* found that 50.9% of this association was mediated by suboptimal socioeconomic status, lifestyle and prepandemic somatic diseases or psychiatric disorders. These findings highlight the role of early life adversities in severe health consequences across the lifespan and call for increased clinical surveillance of people exposed to childhood maltreatment in COVID-19 outbreaks and future pandemics.

## Introduction

Childhood maltreatment, such as sexual, physical and emotional abuse or neglect, is common worldwide, affecting 42.2% of children and adolescents in Europe and 58.4% in North America.^1^ Emerging evidence indicates a role of childhood maltreatment in multiple adverse health outcomes across the lifespan, including psychiatric disorders, cardiovascular diseases, cancers, and some infectious diseases.^2^ For example, children and adolescents who have experienced sexual abuse have an elevated risk of developing genitourinary, respiratory, and ear infections.^3^ Several case reports have further indicated a history of childhood abuse and neglect in fatal systemic Pseudomonas aeruginosa infection,^4^ meningitis,^5^ and recurrent sepsis.^6^

COVID-19 has now resulted in almost 7 million deaths and 100 million hospitalizations worldwide.^7 8^ Among the many assessed risk factors for severe COVID-19 outcomes, including psychosocial factors such as socioeconomic status^9^ and prepandemic history of psychiatric disorders,^10^ childhood maltreatment has largely been overlooked. Childhood maltreatment might impact COVID-19-related morbidity and mortality outcomes through social, behavioral, emotional, and biological pathways.^11 12^ For instance, childhood maltreatment has been associated with significantly higher odds of smoking,^1^ obesity,^1^ cardiometabolic diseases,^1^ psychiatric disorders,^13^ and other chronic somatic diseases in adulthood,^1 2^ all of which are associated with severe COVID-19 outcomes (e.g., hospitalization).^14^ In addition, childhood maltreatment has been associated with immunological dysfunctions,^15^ which may reduce an individual’s capacity to recover from COVID-19.^16^ On the other hand, genetic predisposition to severe COVID-19 outcomes may also play an important role in the risk of COVID-19-related morbidity and mortality. Indeed, several major genetic risk factors for severe COVID-19 outcomes have been identified^17^ and suggested to be associated with an increased risk of severe respiratory failure and death.^18^

However, to our knowledge, only one ecological study has reported a suggestive link between the estimated number of adults with adverse childhood experiences and the number of COVID-19 deaths with county-level data in the US,^19^ leaving the potential role of childhood maltreatment in severe COVID-19 outcomes using individual-level data unravelled. To this end, leveraging the large population-based UK Biobank cohort with prepandemic data on childhood maltreatment, we aimed to explore the association between childhood maltreatment and severe COVID-19 outcomes, i.e., hospitalization or death due to COVID-19, as well as to elucidate the underlying mechanisms and potential role of genetic predisposition to severe COVID-19 outcomes in this association.

## Methods

### Study population and design

We used data from the prospective UK Biobank cohort, which recruited more than 500,000 participants aged 40–69 years from England, Scotland, and Wales between 2006 and 2010. At baseline, participants answered questions on demographic, socioeconomic, lifestyle and health-related factors and provided biological samples for genetic studies.^20^ Then, 339,092 participants who agreed to be contacted again were invited to complete online mental health questionnaires during 2016 and 2017, including a retrospective measure of childhood maltreatment. Of the invited participants, 46.4% (n=157,366) responded to this online measure. Despite those respondents being of higher average socioeconomic status, the reported mental health problems are comparable to the population prevalence estimates for the corresponding age group.^21^

Health-related outcomes (e.g., diagnosis, hospitalization and death) for the participants were obtained periodically through linkage with multiple national datasets. Specifically, hospital inpatient data were obtained from Hospital Episode Statistics for England (from 1997 to September 30^th^, 2021), the Scottish Morbidity Record for Scotland (from 1981 to July 31^st^, 2021), and the Patient Episode Database for Wales (from 1998 to February 28^th^, 2018). Mortality data were obtained from National Health Service (NHS) Digital for England as well as Wales (from 2006 to September 30^th^, 2021) and NHS Central Register for Scotland (from 2006 to October 31^st^, 2021). Records of COVID-19 test results (by RT-PCR of nose/throat swab samples) were obtained through linkage to Public Health England (i.e., PHE, from March 16^th^, 2020 to September 30^th^, 2021), Public Health Scotland (i.e., PHS, from March 16^th^, 2020 to August 31^st^, 2021), and the Secure Anonymised Information Linkage (SAIL) databank (from March 16^th^, 2020 to August 31st, 2021).

In the present study, we included 151,427 participants with information on childhood maltreatment who were alive on January 31^st^ 2020 (i.e., first confirmed COVID-19 cases in the UK) in the analysis. When exploring potential effect modification by genetic predisposition to severe COVID-19 outcomes, we excluded participants of nonwhite British ancestry (n=4,764) or without eligible genotyping data (i.e., genotyping rate < 99%, abnormal heterozygosity level, or kinship coefficient ≥ 0.0884, n=28,870),^22^ leaving 117,793 participants in this analysis. Supplementary Figure 1 shows details of the study population.

### Childhood maltreatment

Childhood maltreatment was measured using the validated short version of the Childhood Trauma Questionnaire (CTQ-5).^23^ It consists of five items assessing whether and how often individuals were exposed to the following types of maltreatment during childhood: sexual abuse, physical neglect, physical abuse, emotional neglect, and emotional abuse, with response options ranging from ‘0’ (never true) to ‘4’ (very often true). The questions and threshold values to define each type of childhood maltreatment are shown in Supplementary Table 1 based on Glaesmer et al.’s study.^23^ In our study, we generated three types of exposure variables: (1) a binary variable indicating any childhood maltreatment, coded as ‘0’ (no) or ‘1’ (yes); (2) a cumulative variable indicating the number of childhood maltreatment types (range from 0 to 5), which was coded as ‘0’, ‘1’, ‘2’, or ‘≥3’ childhood maltreatment types according to the distribution of the entire study sample; and (3) five binary variables for each type of childhood maltreatment, coded as ‘0’ (no) or ‘1’ (yes).

### COVID-19 outcomes

The main outcome of interest was severe COVID-19 outcomes during the study period (i.e., from January 31^st^, 2020, to October 31^st^, 2021). Specifically, participants with a main diagnosis of COVID-19 (ICD-10: U07.1 or U07.2) in hospital inpatient data or with a cause of death recorded as COVID-19 in death registries were defined as having severe COVID-19 outcomes. A secondary outcome of interest was COVID-19 diagnosis, which was determined through records of positive COVID-19 test results in the PHE, PHS and SAIL databanks from March 16^th^, 2020, to September 30^th^, 2021.

### Genetic predisposition to severe COVID-19 outcomes

We assessed genetic predisposition to severe COVID-19 outcomes by calculating the polygenic risk score (PRS) for COVID-19 hospitalization or death according to summary statistics (Version-5) from the COVID19 Host Genetics Initiative large-scale GWAS including critically ill COVID-19 patients (n=4,792) and the control population (n=1,054,664) among individuals with European ancestry after excluding UK Biobank and 23andme participants.^24^ We calculated the PRS using the Clumping + Thresholding approach^25^ under 10 p-value thresholds (i.e., 5 × 10^-8^, 1 × 10^-7^, 1 × 10^-6^, 1 × 10^-5^, 1 × 10^-4^, 1 × 10^-3^, 0.005, 0.01, 0.05, and 0.1) and validated the association between PRS and severe COVID-19 outcomes in our dataset by fitting logistic regression models adjusting for birth year, sex, genotyping array and top ten ancestry principal components. We selected the PRS with the highest Nagelkerke R^2^ for further analyses (i.e., p threshold=5.00×10^-8^; odds ratio=1.21, 95% CI 1.11-1.32; Nagelkerke R^2^=2.01%; Supplementary Table 2). To avoid inflated Type I error from overfitting, we also performed a principal component (PC) analysis on the set of the 10 PRSs and used the first PRS-PC for sensitivity analyses.^26^ In our dataset, the first PRS-PC for severe COVID-19 outcomes showed a strong association with the severe COVID-19 outcome phenotype (odds ratio=1.22, 95% CI 1.11-1.33; Nagelkerke R^2^=2.01%). More information about the PRS-PC analysis is shown in Supplementary Figure 2.

### Covariates

We considered birth year (<1950, 1950-1959 or ≥1960), sex (female or male), ethnicity (Non-White, White, or Unknown) and recruitment region (Scotland, England or Wales) as potential confounders. We studied four variable clusters as potential mediators: 1) socioeconomic status (i.e., Townsend deprivation index [TDI, lower than median, higher than median, or unknown], annual household income [≤£ 180, £ 18000-£ 30999, £ 31000 - £ 51999, £52000 -£100000, ≥£100000, or unknown] and college education [no, yes, or unknown]); 2) lifestyle factors (i.e., smoking status [never, previous, current, or unknown] and body mass index [BMI, <25 kg/m^2^, 25-29.9 kg/m^2^, ≥30 kg/m^2^, or unknown]); 3) prepandemic somatic diseases (no or yes) and 4) prepandemic psychiatric disorders (no or yes). All potential confounders and mediators were selected because of their suggested associations with both childhood maltreatment and severe COVID-19 outcomes (Supplementary Figure 3).

Specifically, TDI was calculated based on the postcode of participants’ address, representing the deprivation at a population level.^27^ BMI was calculated using kilograms (kg) divided by the square of height in meters (m^2^) using anthropometric data measured at the assessment center at baseline. We calculated the Charlson Comorbidity Index (CCI) based on Deyo’s coding algorithm^28^ using hospital inpatient data (before January 31^st^, 2020), and patients with a CCI ≥ 1 were considered to have prepandemic somatic diseases. Supplementary Table 3 provides more details about the diseases included in the calculation of the CCI. We defined prepandemic psychiatric disorders as any diagnosis of psychiatric disorders (ICD-10: F10-F99) in Hospital Inpatient Data before January 31^st^, 2020.

### Statistical analysis

We performed binomial logistic regression to estimate the association between childhood maltreatment and severe COVID-19 outcomes, as well as COVID-19 diagnosis (i.e., secondary outcome), with the estimates reported as odds ratios (ORs) and 95% confidence intervals (CIs). The basic model (Model 1) was adjusted for demographic factors (i.e., birth year, sex, ethnicity and recruitment region). In a stepwise approach, the four variable clusters of mediators were additionally adjusted to examine whether and to what extent the ORs between childhood maltreatment and severe COVID-19 outcomes were attenuated (Models 2-5). We then conducted a regression-based causal mediation analysis^29^ to estimate the proportion of the mediation effect by the four variable clusters of mediators separately (M1-M4) and combined (M5). Specifically, the outcomes were regressed by the primary exposure variable (i.e., exposure to childhood maltreatment), specific variable cluster of mediators and demographic factors in a binomial logistic regression model. Each mediator was then regressed by exposure and demographic factors in either binomial (e.g., prepandemic psychiatric disorders) or multinomial (e.g., BMI) logistic regression models. The results of the outcome and mediator models were then combined to calculate the proportion of mediation.

To determine the association of specific types of childhood maltreatment with severe COVID-19 outcomes, we ran separate analyses for each of the five childhood maltreatment types. Furthermore, we stratified our analyses by tertile of the PRS or the first PRS-PC for severe COVID-19 outcomes (i.e., low: <1st tertile; intermediate: 1st-2nd tertile; high: >2nd tertile) to examine the potential effect modification by genetic predisposition. The statistical significance of the differences between ORs was assessed by introducing interaction terms (i.e., exposure*effect modifier) in the logistic regression adjusted for birth year, sex, ethnicity, and recruitment region, and we obtained a p value to indicate statistical significance through the Wald test.

In sensitivity analyses, we first restricted the analysis of the association between childhood maltreatment and severe COVID-19 outcomes on individuals with COVID-19 diagnosis to address the potential impact of varying risk to COVID-19 across populations. Then, to address the potential impact of COVID-19 vaccination, which started on December 8^th^, 2020, in the UK,^30^ we reran the main analysis by redefining the study period from January 31^st^, 2020, to December 8^th^, 2020 (i.e., before vaccination roll out). Additionally, given the difference in data coverage across registries (e.g., Hospital Inpatient Data and Death registries) and recruitment regions (i.e., England, Scotland, and Wales), we repeated the main analysis by excluding participants registered in Wales as well as by redefining the study period from January 31^st^, 2020 to July 31^st^, 2021. Finally, we performed separate analyses for hospitalization and death due to COVID-19, in addition to considering them as one group.

All analyses were completed using R (version 4.0) and Plink (version 1.9), and a two-tailed test with p < 0.05 was considered statistically significant.

### Patient and public involvement

Patients or the public were not involved in the design, or conduct, or reporting, dissemination plans of our research.

## Results

Of 151,427 participants included in the present study, 56.5% were female, and the mean (SD) age at the start of the pandemic was 67.7 (7.72) years. Nearly one-third (n=50,441) of the participants reported at least one type of childhood maltreatment, and emotional neglect (22.2%) was the most commonly reported type, while physical neglect (5.6%) was the least commonly reported type (Supplementary Table 1). Compared with unexposed individuals, those who were exposed to childhood maltreatment tended to have a lower level of education and annual household income. They were also more likely to be younger, female, obese (i.e., ≥30 kg/m^2^), and with prepandemic somatic diseases as well as psychiatric disorders (Table 1).

**Table 1.** Characteristics of the study population.

|  | Childhood maltreatment |  | Overall<br>(n=151427) |
| --- | --- | --- | --- |
|  | No<br>(n=100986) | Yes<br>(n=50441) |  |
| Age at measure of childhood maltreatment |  |  |  |
| Mean (SD) | 64.1 (7.69) | 63.3 (7.75) | 63.8 (7.72) |
| Median [Min, Max] | 65.0 [46.0, 81.0] | 64.0 [46.0, 80.0] | 65.0 [46.0, 81.0] |
| Age at start of the pandemic (i.e., 2020) |  |  |  |
| Mean (SD) | 68.0 (7.69) | 67.2 (7.75) | 67.7 (7.72) |
| Median [Min, Max] | 69.0 [50.0, 84.0] | 68.0 [50.0, 84.0] | 69.0 [50.0, 84.0] |
| Birth year |  |  |  |
| <1950 | 44042 (43.6%) | 19711 (39.1%) | 63753 (42.1%) |
| 1950-1959 | 36316 (36.0%) | 18997 (37.7%) | 55313 (36.5%) |
| ≥1960 | 20628 (20.4%) | 11733 (23.3%) | 32361 (21.4%) |
| Sex |  |  |  |
| Female | 55882 (55.3%) | 29709 (58.9%) | 85591 (56.5%) |
| Male | 45104 (44.7%) | 20732 (41.1%) | 65836 (43.5%) |
| Ethnicity |  |  |  |
| Non-White | 2013 (2.0%) | 2250 (4.5%) | 4263 (2.8%) |
| White | 98688 (97.7%) | 47975 (95.1%) | 146663 (96.9%) |
| Unknown | 285 (0.3%) | 216 (0.4%) | 501 (0.3%) |
| Recruitment region |  |  |  |
| Scotland | 7019 (7.0%) | 3137 (6.2%) | 10156 (6.7%) |
| Wales | 3771 (3.7%) | 1795 (3.6%) | 5566 (3.7%) |
| England | 90196 (89.3%) | 45509 (90.2%) | 135705 (89.6%) |
| Townsend deprivation index |  |  |  |
| Lower than median (< -2.43) | 52826 (52.3%) | 22825 (45.3%) | 75651 (50.0%) |
| Higher than median (≥ -2.43) | 48041 (47.6%) | 27540 (54.6%) | 75581 (49.9%) |
| Unknown | 119 (0.1%) | 76 (0.2%) | 195 (0.1%) |
| College education |  |  |  |
| No | 46757 (46.3%) | 24230 (48.0%) | 70987 (46.9%) |
| Yes | 47047 (46.6%) | 21806 (43.2%) | 68853 (45.5%) |
| Unknown | 7182 (7.1%) | 4405 (8.7%) | 11587 (7.7%) |
| Annual household income |  |  |  |
| ≤ £ 18000 | 10924 (10.8%) | 7478 (14.8%) | 18402 (12.2%) |
| £ 18000- £ 30999 | 20838 (20.6%) | 10886 (21.6%) | 31724 (21.0%) |
| £ 31000- £ 51999 | 26403 (26.1%) | 13223 (26.2%) | 39626 (26.2%) |
| £ 52000- £ 100000 | 24926 (24.7%) | 10946 (21.7%) | 35872 (23.7%) |
| ≥ £ 100000 | 7812 (7.7%) | 3195 (6.3%) | 11007 (7.3%) |
| Unknown | 10083 (10.0%) | 4713 (9.3%) | 14796 (9.8%) |
| Smoking status |  |  |  |
| Never | 61041 (60.4%) | 26272 (52.1%) | 87313 (57.7%) |
| Previous | 33519 (33.2%) | 19440 (38.5%) | 52959 (35.0%) |
| Current | 6215 (6.2%) | 4589 (9.1%) | 10804 (7.1%) |
| Unknown | 211 (0.2%) | 140 (0.3%) | 351 (0.2%) |
| Body mass index, kg/m<sup>2</sup> <sup>a</sup> |  |  |  |
| <25 | 40352 (40.0%) | 18488 (36.7%) | 58840 (38.9%) |
| 25-29.9 | 41925 (41.5%) | 20621 (40.9%) | 62546 (41.3%) |
| ≥30 | 18477 (18.3%) | 11191 (22.2%) | 29668 (19.6%) |
| Unknown | 232 (0.2%) | 141 (0.3%) | 373 (0.2%) |
| Prepandemic somatic diseases <sup>b</sup> |  |  |  |
| No | 73538 (72.8%) | 35050 (69.5%) | 108588 (71.7%) |
| Yes | 27448 (27.2%) | 15391 (30.5%) | 42839 (28.3%) |
|  | No<br>(n=100986) | Yes<br>(n=50441) |  |
| **Prepandemic psychiatric disorders** |  |  |  |
| No | 94307 (93.4%) | 44493 (88.2%) | 138800 (91.7%) |
| Yes | 6679 (6.6%) | 5948 (11.8%) | 12627 (8.3%) |

a. The body mass index was calculated using weight kilograms (kg) by the square of height in meters (m<sup>2</sup>), using anthropometric data measured at the assessment center at baseline.
b. We calculated the Charlson Comorbidity Index using hospital inpatient data (before January 31<sup>st</sup>, 2020), and patients with a CCI $\geq 1$ were considered to have prepandemic somatic diseases.

A total of 606 individuals were hospitalized (n=542) or died (n=64) as a result of COVID-19 during the study period. We observed increased odds of severe COVID-19 outcomes among patients exposed to any childhood maltreatment (OR=1.54 [95% CI 1.31-1.81]; Table 2) when compared with unexposed individuals in the basic model (Model 1). The association was amplified in a graded manner by the cumulative number of childhood maltreatment types (*p for trend* < 0.01). Specifically, those who experienced three or more childhood maltreatment types had the highest odds of severe COVID-19 outcomes (2.32 [1.73-3.05]), followed by those who experienced two (1.62 [1.22-2.10]) or one (1.33 [1.09-1.62]) type. Inclusion of potential mediators in the models attenuated the magnitude of the association, although ORs remained statistically significantly higher than one in the fully adjusted model (Model 5) among individuals with any childhood maltreatment (1.26 [1.07-1.48]) and those who experienced three or more types of childhood maltreatment (1.50 [1.11-1.98]). Of the five types of childhood maltreatment, physical neglect yielded the strongest association with severe COVID-19 outcomes in the basic model (Model 1, 2.04 [1.57-2.62]; Figure 1) as well as in the model adjusted for all variable clusters of mediators (Model 5, 1.52 [1.16-1.96]), although the difference was not statistically significant (*p for difference* > 0.05).

**Figure 1.**
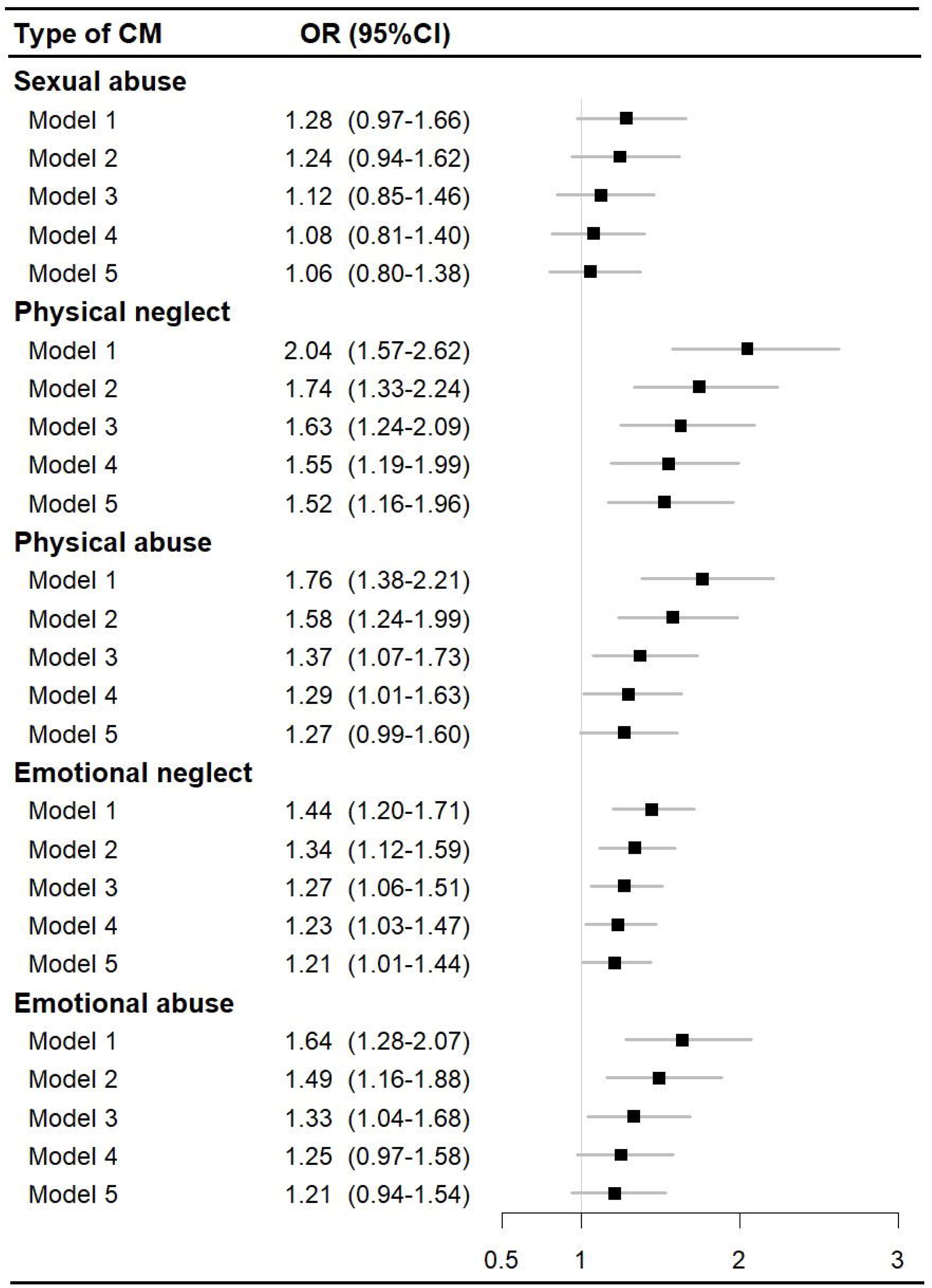
Association between childhood maltreatment (CM) and severe COVID-19 outcomes (i.e., hospitalization or death due to COVID-19) by types of childhood maltreatment. Note: Model 1: Adjusted for demographic factors (birth year, sex, ethnicity, and recruitment region); Model 2: Model 1 and additionally adjusted for socioeconomic factors (townsend deprivation index, college education, and annual household income); Model 3: Model 2 and additionally adjusted for lifestyle-related factors (smoking status and body mass index); Model 4: Model 3 and additionally adjusted for prepandemic somatic diseases (Charlson Comorbidity Index ≥1, before January 31st, 2020); Model 5: Model 4 and additionally adjusted for prepandemic psychiatric disorders (ICD-10: F10-F99; before January 31st, 2020).

**Table 2.**
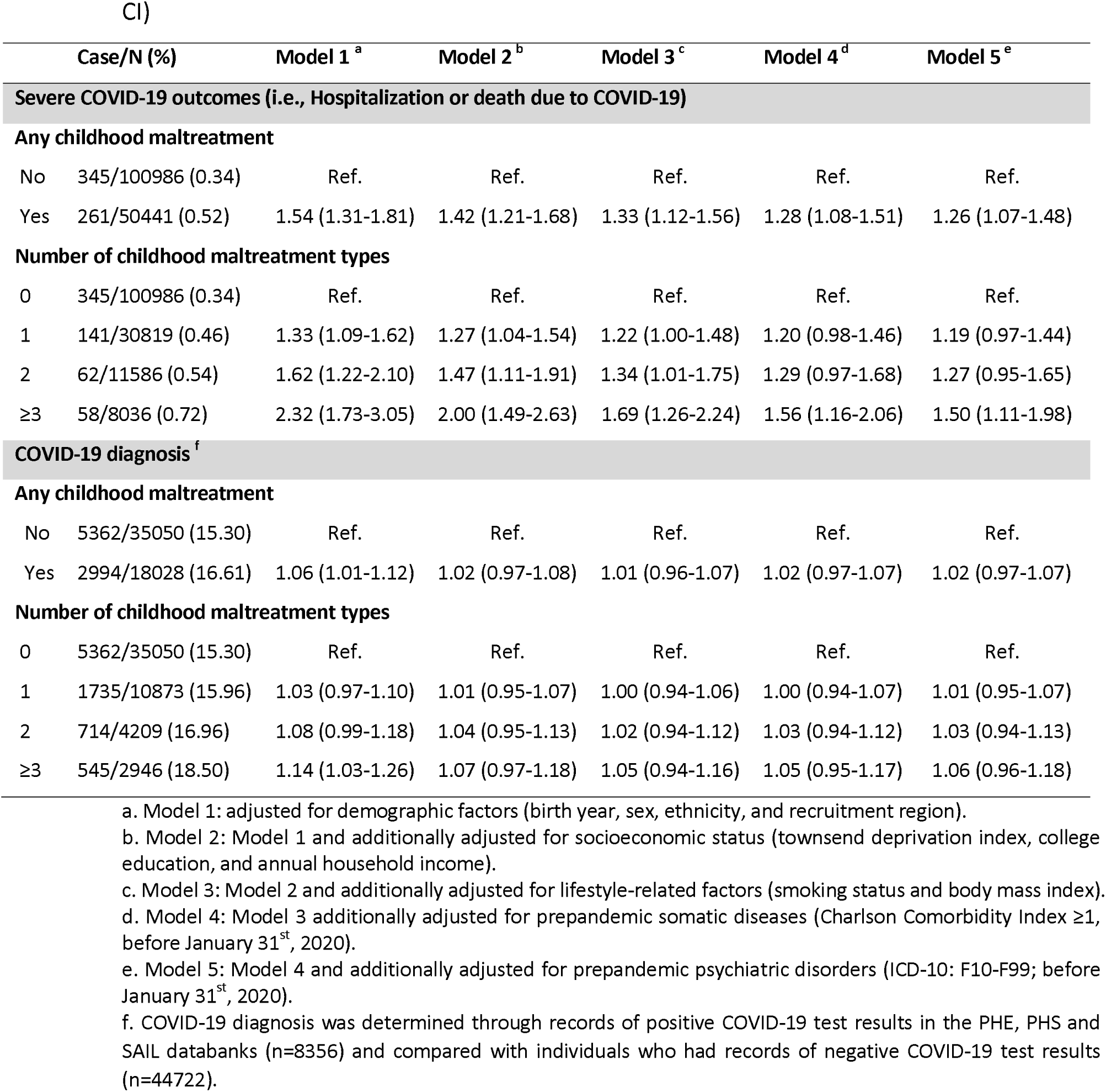
Association between childhood maltreatment and COVID-19 outcomes (OR and 95% CI)

The majority of the association between childhood maltreatment and severe COVID-19 outcomes was mediated through lifestyle factors (27.8%; Figure 2), followed by socioeconomic factors (20.2%), prepandemic somatic diseases (17.4%) and psychiatric disorders (16.6%). In total, 50.9% of the association was mediated by all studied mediators and ranged from 49.5% (after physical neglect) to 79.0% (after sexual abuse) across different types of childhood maltreatment.

**Figure 2.**
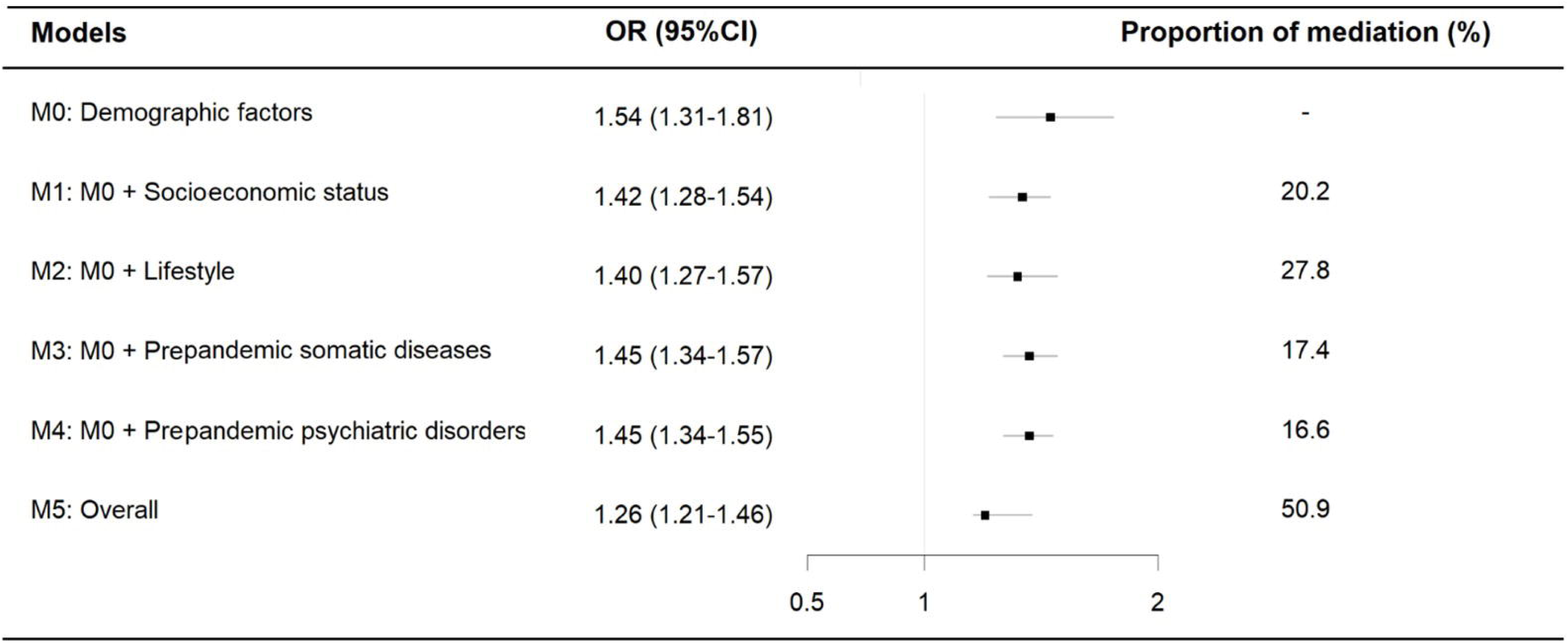
Mediating role of socioeconomic status, lifestyle, prepandemic somatic diseases and psychiatric disorders on the associations between childhood maltreatment and severe COVID-19 outcomes (i.e., hospitalization or death due to COVID-19). Note: M0: adjusted for demographic factors (birth year, sex, ethnicity, and recruitment region); M1: M0 and additionally adjusted for socioeconomic status (townsend deprivation index, college education, and annual household income); M2: M0 and additionally adjusted for lifestyle-related factors (smoking status and body mass index); M3: M0 and additionally adjusted for prepandemic somatic diseases (Charlson Comorbidity Index ≥1, before January 31st, 2020); M4: M0 and additionally adjusted for prepandemic psychiatric disorders (ICD-10: F10-F99; before January 31st, 2020); M5: M0 and additionally adjusted for socioeconomic status, lifestyle, and prepandemic somatic diseases psychiatric disorders; Proportion of mediation: the proportion of the total effect that is mediated through the specified mediators.

We obtained largely comparable results when stratified by tertiles of PRS for severe COVID-19 outcomes (*p for difference* > 0.05; Figure 3). Specifically, exposure to any childhood maltreatment (ORs 1.41-1.88) and three or more types of childhood maltreatment (ORs 2.14-3.11) was consistently associated with significantly increased odds of severe COVID-19 outcomes, regardless of PRS for severe COVID-19 outcomes. We observed similar results when stratifying by the first PRS-PC for severe COVID-19 outcomes (Supplementary Figure 4).

**Figure 3.**
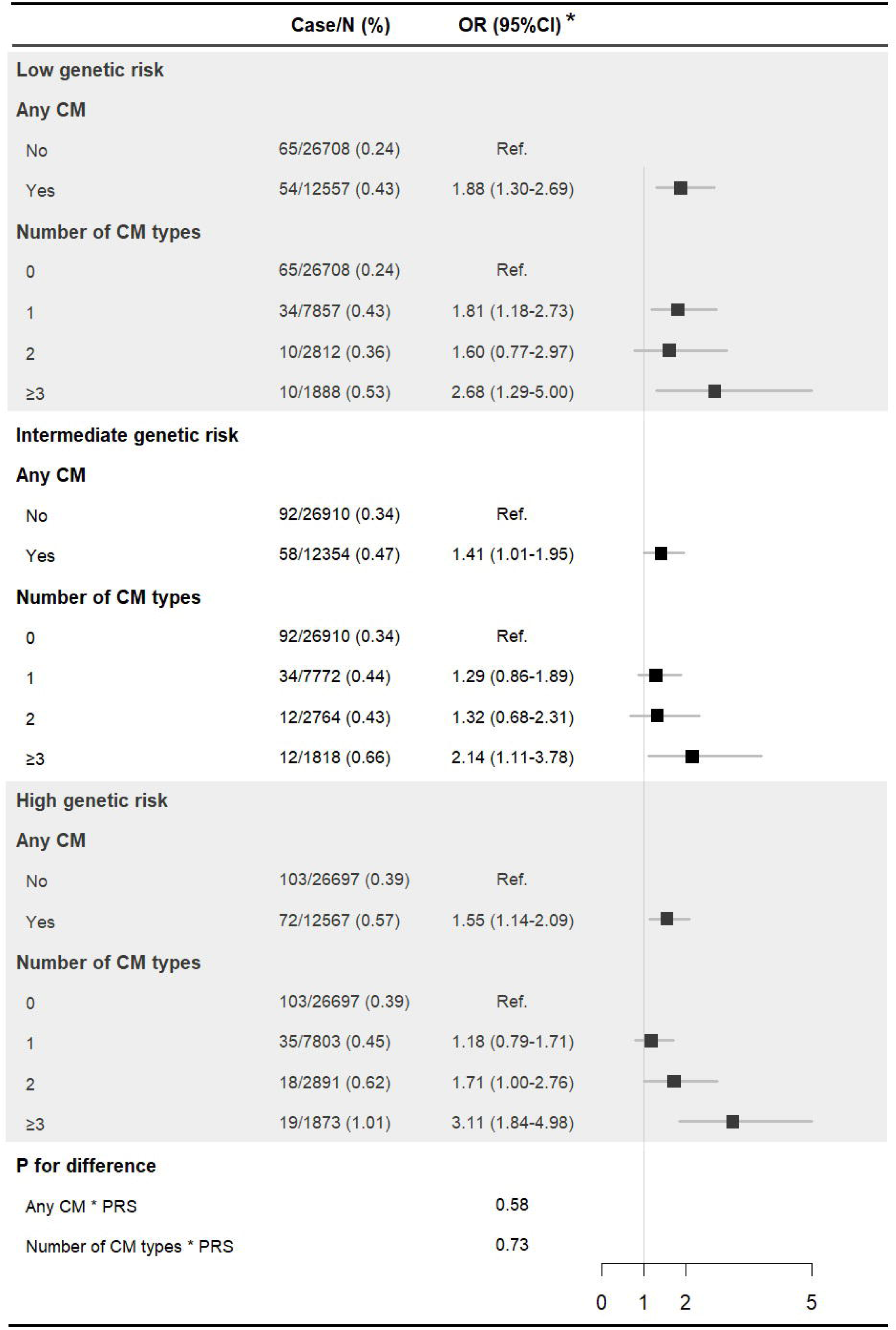
Association between childhood maltreatment (CM) and severe COVID-19 outcomes (i.e., hospitalization or death due to COVID-19) by levels of polygenic risk score (PRS) to severe COVID-19 outcomes. * Adjusted for demographic factors (birth year, sex, ethnicity, and recruitment region).

In the sensitivity analyses, we obtained largely comparable results when restricting our analysis to individuals with a COVID-19 diagnosis (Supplementary Table 4), redefining the study period before vaccination roll out (Supplementary Table 5), excluding participants registered in Wales and redefining the study period from January 31^st^, 2020, to July 31^st^, 2021 (Supplementary Table 6), or after separating hospitalization and death due to COVID-19 as outcomes (Supplementary Table 7).

Finally, in the secondary analysis, we found a weak association between childhood maltreatment and COVID-19 diagnosis (Model 1, 1.06 [1.01-1.12]), which attenuated to null when adding potential mediators to the model (Models 2-5; Table 2).

## Discussion

The findings of this cohort study with prepandemic data on childhood maltreatment suggest a robust dose-response association between childhood maltreatment and severe COVID-19 outcomes. Of the different types of childhood maltreatment, physical neglect in childhood yielded the strongest association with severe COVID-19 outcomes. The associations were partly mediated by suboptimal socioeconomic status, lifestyle and prepandemic psychiatric disorders or other chronic somatic conditions and were not modified by genetic predisposition to severe COVID-19 outcomes.

To our knowledge, this is the first study to explore the association between childhood maltreatment and severe COVID-19 outcomes with individual-level data. Our findings gain support from a previous ecological study^19^ reporting associations between adverse childhood experiences and county-level mortality, as well as a link between early adversity and susceptibility to infectious diseases.^3 5 31^ Our findings also gain support from other studies suggesting an increased risk of illness among women with HIV who were previously exposed to sexual or physical abuse.^32^ In contrast, we observed a weak or no association between childhood maltreatment and COVID-19 diagnosis, which may be due to the widespread transmission of COVID-19 across the population.^7^

Interestingly, we found that physical neglect, measured as being subjected to neglect for necessary medical care during childhood, yielded a somewhat stronger association with severe COVID-19 outcomes than other types of childhood maltreatment, including assault. Indeed, physical neglect has previously been identified as a strong predictor of long-term physical outcomes, including test-identified sexually transmitted infections, diabetes and lung disease.^33 34^ Although the mechanisms behind this finding remain unclear, physical neglect often involves chronic situations that may have cumulative and lasting effects on subsequent health and the propensity to recover from infectious diseases.^35^ In addition, studies have suggested that individuals who experience physical neglect may not receive necessary medical care in childhood and are less likely to seek medical help or treatment in general, contributing to the increased risk of adverse health consequences in adulthood.^36^

We noted that more than half of the association between childhood maltreatment and severe COVID-19 outcomes was mediated by suboptimal socioeconomic status, lifestyle and comorbid psychiatric or other chronic somatic conditions. These results are consistent with previous findings suggesting that childhood maltreatment may increase the risk of health problems in adulthood through multiple factors, including adoption of health-harming behaviors and vulnerabilities to conditions such as obesity and other chronic medical conditions of relevance for COVID-19 severity.^11 37^ We also found the association between childhood maltreatment and severe COVID-19 outcomes to be mediated by psychiatric disorders. Indeed, there is strong evidence for the associations between childhood maltreatment and the risk of psychiatric disorders,^2^ added by our^10^ and more recent findings^38^ indicating a role of preexisting psychiatric disorders in severe COVID-19 outcomes. Of note, among the four studied variable clusters of mediators, lifestyle-related factors appear to have the strongest contribution to the association between childhood maltreatment and severe COVID-19 outcomes; however, there is an established link between lifestyle factors and socioeconomic status,^39^ as well as multiple diseases, including cardiometabolic conditions^40^ and mental disorders.^41^ Therefore, the proportion mediated by each cluster of mediators, as suggested in the causal mediation analysis, is likely confounded by the other mediating clusters. Nonetheless, knowledge on these potential mediating factors could help inform targeted interventions and mitigate the harmful effects of childhood maltreatment on severe health consequences in the COVID-19 outbreak and future pandemics.

As the association between childhood maltreatment and severe COVID-19 outcomes was still robust after controlling for these potential mediators, other biological pathways through which childhood maltreatment may lead to lasting physiological changes, including disruption of inflammatory responses^42^ and hormonal dysregulation,^43^ and contribute to the elevated risk of severe COVID-19 outcomes. For instance, recent evidence suggests that elevated IL-6 and TNF-α levels can predict disease severity and survival in patients with COVID-19.^16^ In addition, our findings indicate that the influence of childhood maltreatment on severe COVID-19 outcomes is not modified by genetic predisposition to such adverse outcomes, suggesting that additional environmental risk factors, such as childhood maltreatment, may play an equal role across groups with varying genetic liability to these adverse outcomes.

### Strengths and limitations of this study

The major strength of our study is the use of a prospective study design, i.e., prepandemic individual data on childhood maltreatment and follow-up data on COVID-19, in a large population-based cohort. This ensures that the measures of childhood maltreatment indeed preceded any severe COVID-19 outcomes and hence minimizes the risk of reverse causality. Additionally, the primary outcome of interest was severe COVID-19 outcomes (i.e., hospitalization or death), and the secondary analysis was restricted to patients tested for COVID-19 (i.e., tested positive vs. tested negative); thus, the influence of surveillance bias should be minor. Moreover, our consideration of genetic predisposition to severe COVID-19 outcomes and a wide range of mediators provides evidence of pathways linking childhood adversities to severe COVID-19 outcomes, with potential relevance for prevention and intervention strategies.

This study also has several limitations to be noted. First, as in most studies on childhood maltreatment, information on childhood maltreatment was recalled by participants in middle or older age rather than captured prospectively (in childhood), which may be liable to underreport^44 45^ and biased by current mental state.^46^ However, to explain the observed results pattern, such measurement error would have to be systematically differential in relation to later severe COVID-19 outcomes. Second, several included mediators (e.g., smoking status, BMI) were only measured once at baseline and might have changed over the 10-year follow-up. Third, the incidence of COVID-19 varied across populations and geographical regions^47^, potentially with a disproportionate size of the population exposed to childhood maltreatment. However, our sensitivity analysis, which was restricted to individuals with a COVID-19 diagnosis or excluded participants registered in Wales and redefined the study period from January 31^st^, 2020, to July 31^st^, 2021, suggested a minimal influence of these factors on the reported associations. Finally, there is evidence of a ‘healthy volunteer’ selection bias of UK participants, and most severe childhood maltreatment cases were probably not included in the cohort,^48^ possibly resulting in underestimation of the studied association. Indeed, the UK Biobank cohort is not representative of the entire UK population in terms of socioeconomic status or lifestyle factors, and only approximately 30% of the UK Biobank participants were included in our analysis; thus, the generalization of our findings should be made with caution.

## Conclusions

In conclusion, our findings suggest that childhood maltreatment, particularly physical neglect, is robustly associated with severe COVID-19 outcomes (i.e., hospitalization or death due to COVID-19). The association was partly mediated by suboptimal socioeconomic status, lifestyle factors and comorbidities and was not modified by genetic predisposition to severe COVID-19 outcomes. These findings highlight the role of early life adversities in severe health consequences across the lifespan and call for increased clinical surveillance of people exposed to childhood maltreatment in COVID-19 outbreaks and future pandemics.

## Ethics statements

### Ethical approval

This study was approved by the ethics review authority (2022-01516-01) in Sweden. The UK Biobank has approval from the North West Multi-Centre Research Ethics Committee as a Research Tissue Bank approval (11/NW/0382).

## Data availability statement

Data from the UK Biobank (http://www.ukbiobank.ac.uk/) are available to all researchers upon making an application.

## Acknowledgments

This research was conducted using the UK Biobank Resource under Application Number 76517. The computations and data handling were partly enabled by resources provided by the National Academic Infrastructure for Supercomputing in Sweden (NAISS) and the Swedish National Infrastructure for Computing (SNIC) at Uppsala Multidisciplinary Center for Advanced Computational Science (UPPMAX) partially funded by the Swedish Research Council through grant agreements no. 2022-06725 and no. 2018-05973.

## Contributors

UAV designed the study. YW performed the phenotypic analysis, and FG performed the genotypic analysis, with support from UAV, TA, KH and FF. YW, FG and UAV drafted the manuscript, and all authors contributed to data interpretation. All authors approved the final manuscript as submitted and agreed to be accountable for all aspects of the work.

