## Supplementary for "Childhood maltreatment and subsequent risk of hospitalization or death due to COVID-19: a cohort study in the UK Biobank"

Supplenmentary Figure 1 Study profile.

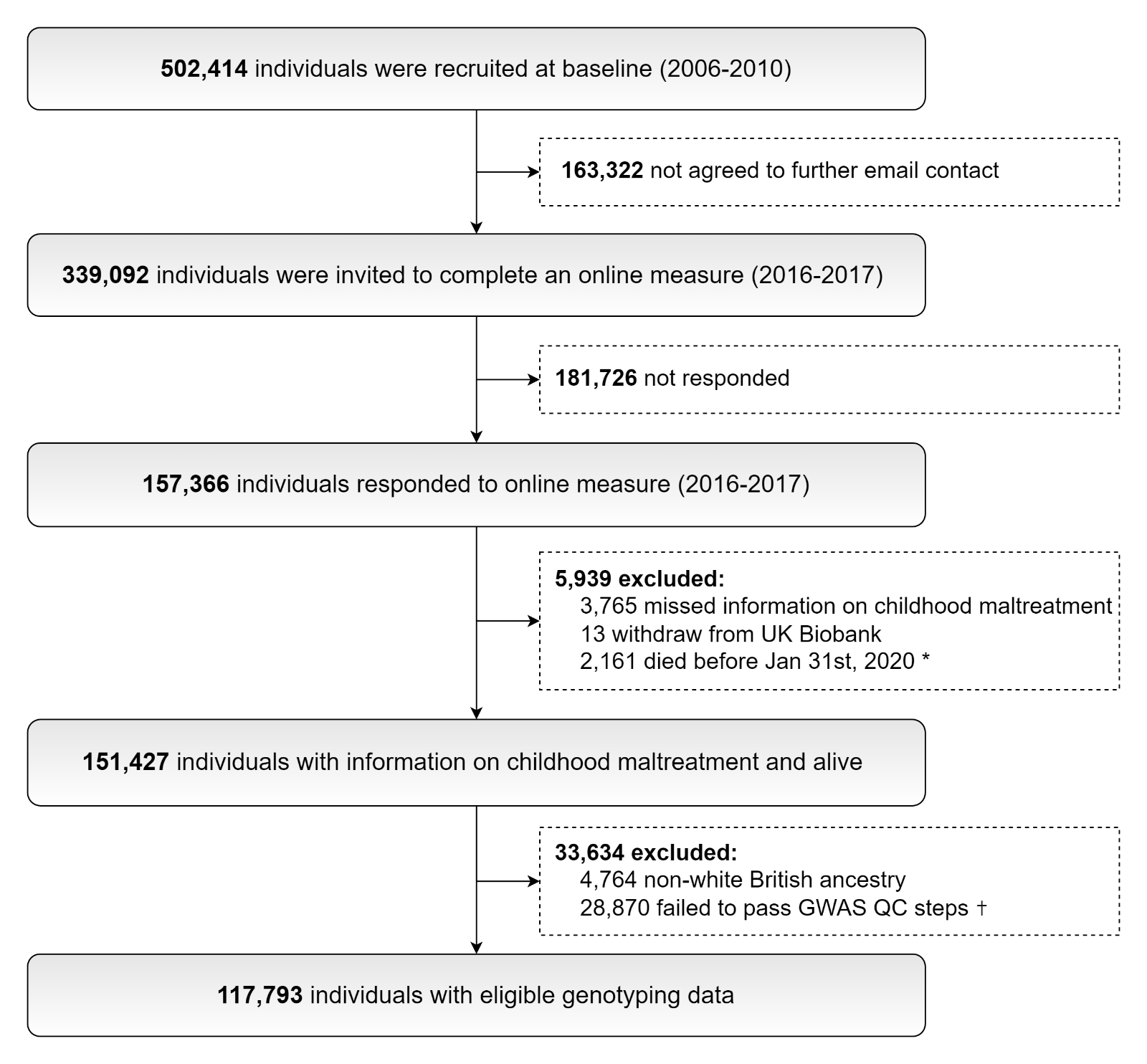

* 1st confirmed COVID-19 cases in the UK

✝ We excluded individuals having genotyping rate <98%, with abnormal heterozygosity level or a kinship coefficient >0.0884.

Supplenmentary Figure 2 Principal compent (PC) on the set of the 10 polygenic risk score (PRS) for severe COVID-19 outcomes.

| A. Correlation of the PRS based on different p-value thresholds in the studied sample (n=117,793). | | |
| --- | --- | --- |
| 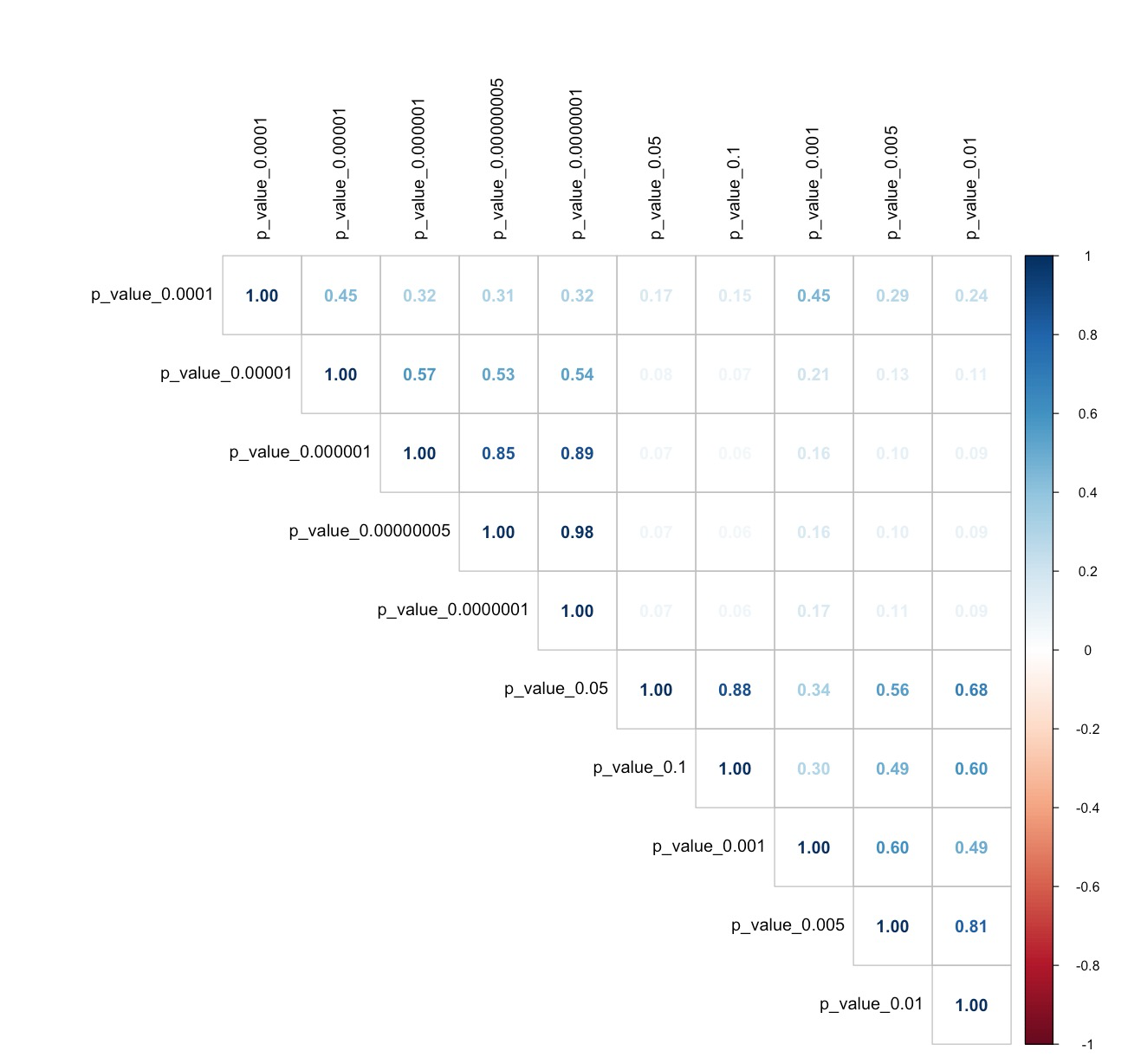 | | |
| B. Percentage explained variance by each PRS-PC. | | C. The loadings in the first PRS-PC at each threshold. |

Explained variance (%)

|  | Percentage explained variance |
| --- | --- |
| PC 1 | 39.9071 |
| PC 2 | 28.588 |
| PC 3 | 10.9053 |
| PC 4 | 7.10995 |
| PC 5 | 4.93557 |
| PC 6 | 3.97442 |
| PC 7 | 1.65209 |
| PC 8 | 1.63088 |
| PC 9 | 1.12171 |
| PC 10 | 0.175029 |

Contributions (%)

|  | PC 1 |
| --- | --- |
| p_value_0.00000005 | 11.8173 |
| p_value_0.0000001 | 12.1617 |
| p_value_0.000001 | 11.4863 |
| p_value_0.00001 | 8.60071 |
| p_value_0.0001 | 8.32478 |
| p_value_0.001 | 9.16807 |
| p_value_0.005 | 10.8024 |
| p_value_0.01 | 10.8114 |
| p_value_0.05 | 9.0382 |
| p_value_0.1 | 7.78914 |

Supplenmentary Figure 3 Proposed causal model with alternative pathways of how childhood maltreatment could influence severe COVID-19 outcomes (i.e., hospitalization or death due to COVID-19), while taking into account the availability of data.

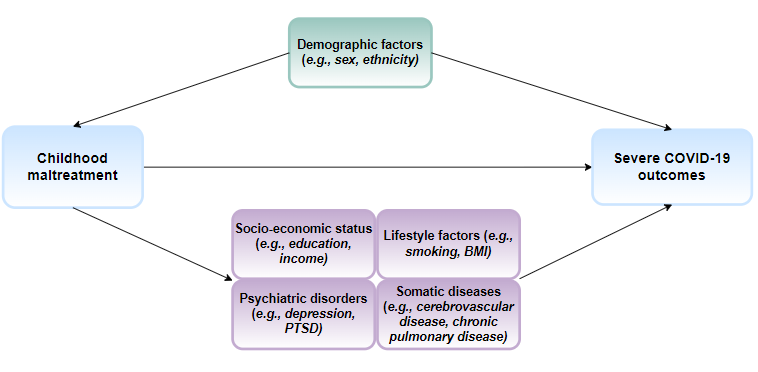

Note, Box in green indicate potential confounders of the association between childhood maltreatment and severe COVID-19 outcomes and boxes in purple indicate potential mediators of the association.

Supplenmentary Figure 4 Association between childhood maltreatment (CM) and severe COVID-19 outcomes (i.e., hospitalization or death due to COVID-19), by levels of first PRS-PC to severe COVID-19 outcomes.

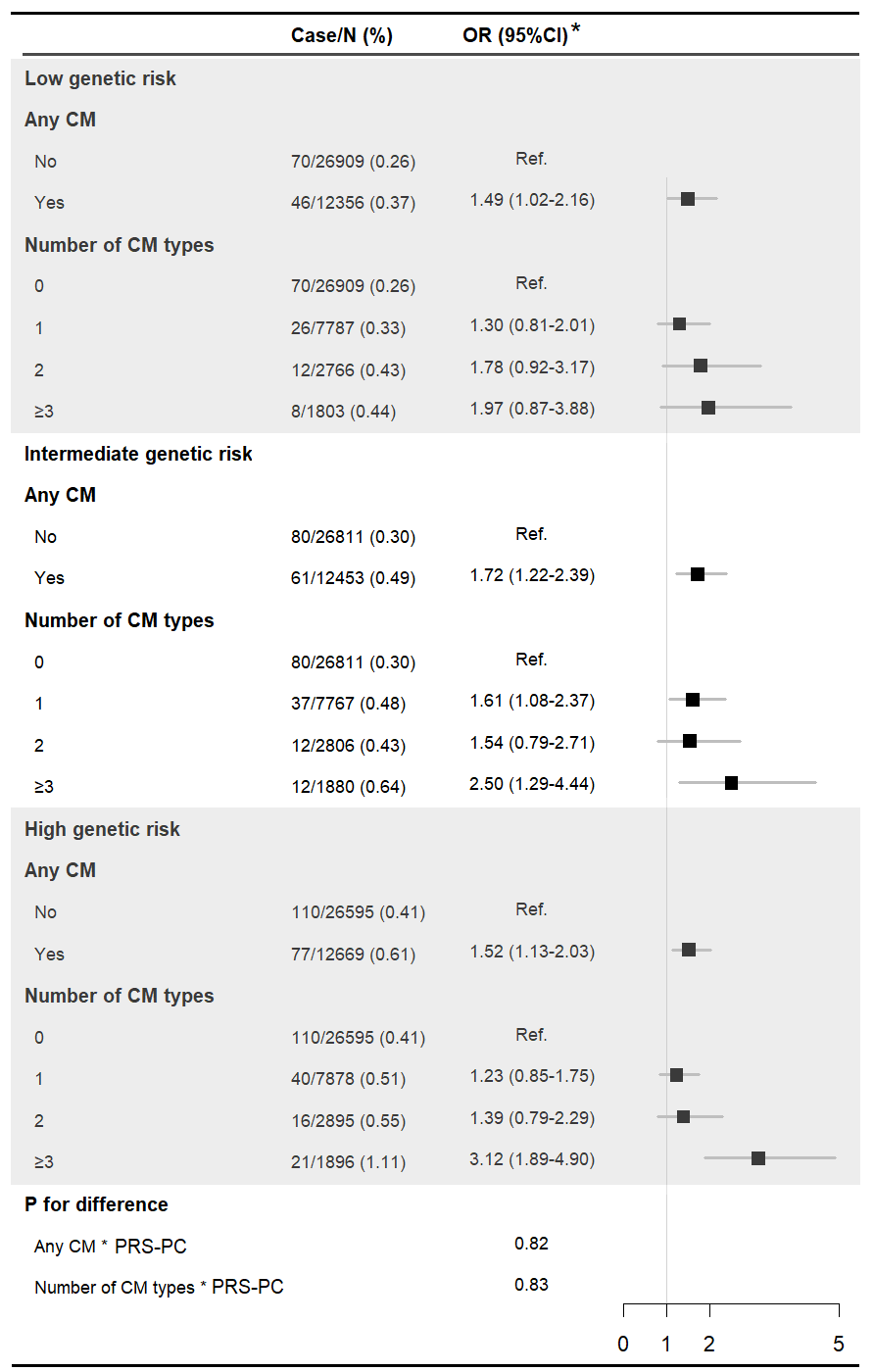

* Adjusted for demographic factors (birth year, sex, ethnicity, and recruitment region).

Supplenmentary Table 1 Participants’ response to the 5 types of childhood maltreatment

| N (%) | Never true | Rarely true | Sometimes true | Often true | Very often true |
| --- | --- | --- | --- | --- | --- |
| Physical abuse: People in my family hit me so hard that it left me with bruises or marks | 123020 (81.2) | 16233 (10.7) | 10009 (6.6) | 1316 (0.9) | 849 (0.6) |
| Emotional abuse: I felt that someone in my family hated me | 128080 (84.6) | 9203 (6.1) | 9895 (6.5) | 2322 (1.5) | 1927 (1.3) |
| Sexual abuse: Someone molested me (sexually) | 138151 (91.2) | 7017 (4.6) | 4854 (3.2) | 766 (0.5) | 639 (0.4) |
| Physical neglect: There was someone to take me to the doctor if I needed it | 3216 (2.1) | 1200 (0.8) | 4051 (2.7) | 15975 (10.6) | 126985 (83.9) |
| Emotional neglect: I felt loved | 2141 (1.4) | 6940 (4.6) | 24473 (16.2) | 38482 (25.4) | 79391 (52.4) |

Note, red fonts indicate the responses that are categorized as exposed to childhood maltreatment.

Supplenmentary Table 2 Associations between polygenic risk scores (PRS, calculated using Clumping + Thresholding approach) for severe COVID-19 outcomes (i.e., hospitalization or death due to COVID-19) at different p value thresholds.

| **P threshold** | **OR (95%CI)** | **Nagelkerke R^2^ (%)** | **p** | **Number of SNPs** |
| --- | --- | --- | --- | --- |
| 5.00×10^-08^ | 1.21 (1.11-1.32) | 2.01 | <0.01 | 8 |
| 1.00×10^-07^ | 1.21 (1.11-1.32) | 2.00 | <0.01 | 9 |
| 1.00×10^-06^ | 1.19 (1.08-1.30) | 1.94 | <0.01 | 12 |
| 1.00×10^-05^ | 1.11 (1.01-1.22) | 1.79 | 0.02 | 37 |
| 1.00×10^-04^ | 1.07 (0.98-1.18) | 1.74 | 0.14 | 184 |
| 0.001 | 1.08 (0.98-1.18) | 1.74 | 0.11 | 1225 |
| 0.005 | 1.10 (1.00-1.21) | 1.77 | 0.05 | 4855 |
| 0.01 | 1.10 (1.00-1.20) | 1.77 | 0.05 | 8854 |
| 0.05 | 1.12 (1.02-1.23) | 1.80 | 0.02 | 34226 |
| 0.1 | 1.13 (1.03-1.24) | 1.82 | 0.01 | 59893 |

Note, GWAS summary statistics for COVID-19 hospitalization were obtained from <https://www.covid19hg.org/results/r5/>. Odds ratio and 95% confidence interval were estimated by logistic regression models, adjusting for birth year, sex, genotyping array, and top 10 ancestry principal components.

Supplenmentary Table 3 Diseases used for calculating Charlson comorbidity index.

| **Disease** | **ICD-10 code** |
| --- | --- |
| Myocardial infarction | I21, I22, I252 |
| Congestive heart failure | I110, I130, I132, I50 |
| Peripheral vascular disease | I70, I71, I731, I738, I739, I771, I790, I792, K551, K558, K559, R02, Z958, Z959 |
| Cerebrovascular disease | I60-I69, G45, G46 |
| Dementia | F00, F01, F02, F03, F051, G30, G311 |
| Chronic pulmonary disease | J40-J47, J60, J61, J62, J63, J64, J65, J66, J67, J684, J70, J841, J920, J961, J982 |
| Connective tissue disease | M05, M06, M30, M315, M32, M33, M34, M351, M353, M360 |
| Ulcer disease | K25, K26, K27, K28 |
| Mild liver disease | B18, K700, K701, K702, K703, K709, K713, K714, K715, K717, K73, K74, K760, K762, K763, K764, K768, K769, |
| Diabetes mellitus | E100, E101, E106, E108, E109, E110, E111, E116, E118, E119, E120, E121, E126, E128, E129, E130, E131, E136, E138, E139, E140, E141, E146, E148, E149, |
| Hemiplegia | G041, G114, G801, G802, G81, G82, G839, G830, G831, G832, G833, G834 |
| Moderate/severe renal disease | I120, I131, N032, N033, N034, N035, N036, N037, N052, N053, N054, N055, N056, N057, N18, N19, N250, Z940, Z992 |
| Diabetes mellitus with chronic complications | E102, E103, E104, E105, E107, E112-E115, E122-E125, E132- E135, E142- E145, E117, E127, E137, E147 |
| Any tumor | C00-C14, C15-C26, C30-C34, C37-C41, C43, C45-C49, C50-C50, C51-C58, C60-C63, C64-C68, C69-C72, C73-C76, C97 |
| Leukemia | C91, C92, C93, C94, C95 |
| Lymphoma | C81, C82, C83, C84, C85, C88, C90, C96 |
| Moderate/severe liver disease | I85, K704, K72, K766 |
| Metastatic solid tumor | C77, C78, C79, C80 |
| AIDS | B20, B21, B22, B23, B24 |

Supplenmentary Table 4 Association between childhood maltreatment and severe COVID-19 outcomes (i.e., hospitalization or death due to COVID-19; OR and 95%CI), restricted analysis to individuals with COVID-19 diagnosis

|  | **Case/N (%)** | **Model 1 ^a^** | **Model 2 ^b^** | **Model 3 ^c^** | **Model 4 ^d^** | **Model 5 ^e^** |
| --- | --- | --- | --- | --- | --- | --- |
| **Any childhood maltreatment** | | | | | | |
| No | 339/5362 (6.32) | Ref. | Ref. | Ref. | Ref. | Ref. |
| Yes | 254/2994 (8.48) | 1.44 (1.21-1.72) | 1.39 (1.16-1.65) | 1.29 (1.08-1.54) | 1.22 (1.02-1.46) | 1.20 (1.00-1.44) |
| **Number of childhood maltreatment types** | | | | | | |
| 0 | 339/5362 (6.32) | Ref. | Ref. | Ref. | Ref. | Ref. |
| 1 | 138/1735 (7.95) | 1.30 (1.05-1.60) | 1.28 (1.03-1.58) | 1.22 (0.98-1.51) | 1.18 (0.95-1.47) | 1.17 (0.94-1.45) |
| 2 | 59/714 (8.26) | 1.42 (1.05-1.90) | 1.33 (0.98-1.78) | 1.24 (0.91-1.66) | 1.17 (0.85-1.57) | 1.15 (0.84-1.55) |
| ≥3 | 57/545 (10.46) | 2.11 (1.54-2.84) | 1.94 (1.41-2.63) | 1.63 (1.18-2.21) | 1.44 (1.04-1.98) | 1.39 (1.00-1.91) |

a. Model 1: adjusted for demographic factors (birth year, sex, ethnicity, and recruitment region).

b. Model 2: Model 1 and additionally adjusted for socioeconomic status (Townsend deprivation index, college education, and annual household income).

c. Model 3: Model 2 and additionally adjusted for lifestyle-related factors (smoking status and body mass index).

d. Model 4: Model 3 and additionally adjusted for pre-pandemic somatic diseases (Charlson Comorbidity Index ≥1, before January 31^st^, 2020).

e. Model 5: Model 4 and additionally adjusted for pre-pandemic psychiatric disorders (ICD-10: F10-F99; before January 31^st^, 2020).

Supplenmentary Table 5 Association between childhood maltreatment and severe COVID-19 outcomes (i.e., hospitalization or death due to COVID-19; OR and 95%CI), re-defining the study period from January 31^st^, 2020 to December 8^th^, 2020 (i.e., before vaccination roll out).

|  | **Case/N (%)** | **Model 1 ^a^** | **Model 2 ^b^** | **Model 3 ^c^** | **Model 4 ^d^** | **Model 5 ^e^** |
| --- | --- | --- | --- | --- | --- | --- |
| **Any childhood maltreatment** | | | | | | |
| No | 169/100986 (0.17) | Ref. | Ref. | Ref. | Ref. | Ref. |
| Yes | 132/50441 (0.26) | 1.59 (1.26-2.00) | 1.46 (1.16-1.84) | 1.36 (1.08-1.71) | 1.31 (1.04-1.65) | 1.28 (1.01-1.61) |
| **Number of childhood maltreatment types** | | | | | | |
| 0 | 169/100986 (0.17) | Ref. | Ref. | Ref. | Ref. | Ref. |
| 1 | 72/30819 (0.23) | 1.39 (1.05-1.83) | 1.32 (1.00-1.73) | 1.27 (0.96-1.67) | 1.25 (0.94-1.64) | 1.23 (0.93-1.62) |
| 2 | 31/11586 (0.27) | 1.65 (1.10-2.38) | 1.49 (0.99-2.16) | 1.36 (0.90-1.97) | 1.31 (0.87-1.89) | 1.27 (0.85-1.84) |
| ≥3 | 29/8036 (0.36) | 2.37 (1.56-3.48) | 2.02 (1.32-2.97) | 1.69 (1.10-2.50) | 1.54 (1.01-2.28) | 1.45 (0.94-2.15) |

a. Model 1: adjusted for demographic factors (birth year, sex, ethnicity, and recruitment region).

b. Model 2: Model 1 and additionally adjusted for socioeconomic status (Townsend deprivation index, college education, and annual household income).

c. Model 3: Model 2 and additionally adjusted for lifestyle-related factors (smoking status and body mass index).

d. Model 4: Model 3 and additionally adjusted for pre-pandemic somatic diseases (Charlson Comorbidity Index ≥1, before January 31^st^, 2020).

e. Model 5: Model 4 and additionally adjusted for pre-pandemic psychiatric disorders (ICD-10: F10-F99; before January 31^st^, 2020).

Supplenmentary Table 6 Association between childhood maltreatment and severe COVID-19 outcomes (i.e., hospitalization or death due to COVID-19; OR and 95%CI), excluding participants registered in Wales as well as re-defining the study period from January 31^st^, 2020 to July 31^st^, 2021.

|  | **Case/N (%)** | **Model 1 ^a^** | **Model 2 ^b^** | **Model 3 ^c^** | **Model 4 ^d^** | **Model 5 ^e^** |
| --- | --- | --- | --- | --- | --- | --- |
| **Any childhood maltreatment** | | | | | | |
| No | 311/97215 (0.32) | Ref. | Ref. | Ref. | Ref. | Ref. |
| Yes | 240/48646 (0.49) | 1.56 (1.31-1.85) | 1.44 (1.21-1.71) | 1.34 (1.13-1.59) | 1.30 (1.09-1.54) | 1.27 (1.07-1.51) |
| **Number of childhood maltreatment types** | | | | | | |
| 0 | 311/97215 (0.32) | Ref. | Ref. | Ref. | Ref. | Ref. |
| 1 | 127/29668 (0.43) | 1.33 (1.08-1.63) | 1.27 (1.02-1.55) | 1.22 (0.98-1.49) | 1.19 (0.97-1.47) | 1.18 (0.96-1.45) |
| 2 | 56/11218 (0.50) | 1.60 (1.19-2.12) | 1.45 (1.08-1.92) | 1.33 (0.98-1.75) | 1.28 (0.95-1.69) | 1.26 (0.93-1.66) |
| ≥3 | 57/7760 (0.73) | 2.49 (1.85-3.29) | 2.14 (1.59-2.84) | 1.81 (1.34-2.41) | 1.67 (1.24-2.23) | 1.61 (1.19-2.15) |

a. Model 1: adjusted for demographic factors (birth year, sex, ethnicity, and recruitment region).

b. Model 2: Model 1 and additionally adjusted for socioeconomic status (Townsend deprivation index, college education, and annual household income).

c. Model 3: Model 2 and additionally adjusted for lifestyle-related factors (smoking status and body mass index).

d. Model 4: Model 3 and additionally adjusted for pre-pandemic somatic diseases (Charlson Comorbidity Index ≥1, before January 31^st^, 2020).

e. Model 5: Model 4 and additionally adjusted for pre-pandemic psychiatric disorders (ICD-10: F10-F99; before January 31^st^, 2020).

Supplenmentary Table 7 Association between any childhood maltreatment and severe COVID-19 outcomes, separating for hospitalization and death due to COVID-19 (OR and 95%CI).

|  | **Case/N (%)** | **Model 1 ^a^** | **Model 2 ^b^** | **Model 3 ^c^** | **Model 4 ^d^** | **Model 5 ^e^** |
| --- | --- | --- | --- | --- | --- | --- |
| **Hospitalization due to COVID-19** | | | | | | |
| **Any childhood maltreatment** | | | | | | |
| No | 307/100948 (0.30) | Ref. | Ref. | Ref. | Ref. | Ref. |
| Yes | 235/50415 (0.47) | 1.54 (1.30-1.83) | 1.44 (1.21-1.71) | 1.34 (1.12-1.59) | 1.29 (1.08-1.53) | 1.27 (1.06-1.51) |
| **Death due to COVID-19** | | | | | | |
| **Any childhood maltreatment** | | | | | | |
| No | 38/100679 (0.04) | Ref. | Ref. | Ref. | Ref. | Ref. |
| Yes | 26/50206 (0.05) | 1.52 (0.91-2.50) | 1.37 (0.82-2.26) | 1.34 (0.80-2.20) | 1.29 (0.77-2.14) | 1.26 (0.75-2.09) |

a. Model 1: adjusted for demographic factors (birth year, sex, ethnicity, and recruitment region).

b. Model 2: Model 1 and additionally adjusted for socioeconomic status (townsend deprivation index, college education, and annual household income).

c. Model 3: Model 2 and additionally adjusted for lifestyle-related factors (smoking status and body mass index).

d. Model 4: Model 3 and additionally adjusted for pre-pandemic somatic diseases (Charlson Comorbidity Index ≥1, before January 31^st^, 2020).

e. Model 5: Model 4 and additionally adjusted for pre-pandemic psychiatric disorders (ICD-10: F10-F99; before January 31^st^, 2020).
